# Socio-economic deprivation and cancer incidence in England: Quantifying the role of smoking

**DOI:** 10.1101/2022.02.11.22270853

**Authors:** Nick W. S. Payne, Katrina F. Brown, Christine Delon, Yannis Kotrotsios, Isabelle Soerjomataram, Jon Shelton

## Abstract

**Background:** More deprived populations typically experience higher cancer incidence rates and smoking prevalence compared to less deprived populations. We calculated the proportion of cancer cases attributable to smoking by socio-economic deprivation in England and estimated the impact smoking has on the deprivation gap for cancer incidence.

**Methods:** Data for cancer incidence (2013-2017), smoking prevalence (2003-2007) and population estimates (2013-2017) were split by sex, age-group and deprivation quintile. Relative risk estimates from meta-analyses were used to estimate the population attributable fraction (PAF) for 15 cancer types associated with smoking. The deprivation gap was calculated using age-specific incidence rates by deprivation quintile.

**Results:** Smoking-related cancer PAFs in England are 2.2 times larger in the most deprived quintile compared to the least deprived quintile (from 9.7% to 21.1%). If everyone had the same smoking prevalence as the least deprived quintile, 20% of the deprivation gap in cancer incidence could have been prevented. If nobody smoked, 61% of the deprivation gap could have been prevented.

**Conclusions:** The majority of the deprivation gap in cancer incidence could have been prevented in England between 2013-2017 if nobody had smoked. Policy makers should ensure that tobacco control policies reduce overall smoking prevalence by tackling smoking inequalities.

## Background

Smoking is the main cause of preventable cancer and death in the UK.^1,2^ In England, smoking accounted for 15% (around 44,000 cases) of all cancer cases in 2015.^1^ Smoking causes at least 15 different types of cancer, and the proportion of cases caused by smoking varies greatly by cancer type, ranging from 0.3% for ovarian cancer to 72% for lung cancer in England.

Cancer incidence varies by socio-economic position across the UK.^3,4,5,6^ For example, cancer incidence rates for all cancers combined in England are 17% higher in the most deprived quintile compared to the least.^7^

The majority of cancer types’ incidence rates are positively associated with deprivation in England, leading to an estimated 27,200 deprivation-associated cancer cases each year.^3^ Many of the cancer types associated with deprivation are also associated with smoking.^3,8^

A clear socio-economic divide is observed for adult smoking prevalence in the UK.^9^ In England, smoking prevalence is around 2.5 times higher in the most deprived group compared to the least deprived group.^9,10^ In line with this, previous studies in France and Australia have reported that more deprived populations had a higher burden of cancer incidence attributable to smoking.^11,12^ These studies also investigated the impact of the removal of smoking inequalities, which estimated that 7-13% of all cancers caused by smoking in men and 8-9% in women, could be prevented if everyone smoked like the least deprived quintile.

We aimed to estimate the proportion of cancer cases attributable to smoking by socio-economic position in England. Additionally, we estimated what proportion of the observed deprivation gap in cancer incidence in England could have been prevented if: 1) everyone had the same smoking prevalence as the least deprived group; 2) nobody smoked.

## Methods

### Cancer Types

To calculate the proportion of cancer cases attributable to smoking we included 15 cancer types which have ‘sufficient’ evidence of a causal association with smoking based on the International Agency for Research on Cancer (IARC) Monograph^8^: oral cavity, pharynx, nasopharynx, larynx, oesophagus, stomach, colorectal, liver, pancreas, lung, cervix uteri, kidney, bladder, ovarian (mucinous) and acute myeloid leukaemia (see Supplementary Material A for International Classification of Diseases version 10 codes). These cancers contribute to 44% (around 134,300 cases) of the total cancer incidence in England every year (2013-2017). We will refer to these cancer types as ‘smoking-related cancers’.

Only cancer types positively associated with deprivation – defined as having significantly higher age-standardised incidence rates in the most deprived quintile compared to the least deprived – between 2013 and 2017 in England were included for calculation of the observed deprivation gap in cancer incidence^3^: head and neck (oral cavity, salivary glands, pharynx, nasopharynx, larynx, nasal cavity and middle ear, accessory sinuses), oesophagus, stomach, colorectal, liver, pancreas, lung, cervix uteri, kidney, bladder, small intestine, anal, gallbladder, vulva, vagina, uterus, penis, Hodgkin lymphoma and cancer of unknown primary (see Supplementary Material A for ICD-10 codes). These cancers contribute to 50% (around 154,000 cases) of the total cancer incidence in England every year (2013-2017). We will refer to these cancer types as ‘deprivation-related cancer types’.

## Data sources

Cancer incidence for England between 2013 and 2017 was provided by Public Health England and population estimates between 2013 and 2017 were provided by the Office for National Statistics. Each data set was split by sex, 5-year age band and quintiles of the Income domain from the Index of Multiple Deprivation 2015 (IMD 2015). The Income domain of the IMD is a relative, local-level measure of deprivation based on the proportion of the population in that area estimated to experience deprivation because of low income. The cancer data was additionally split by ICD-10 3-digit code, or International Classification of Diseases for Oncology version 3 (ICD-O-3) code (e.g. mucinous ovarian, oesophageal adenocarcinoma and oesophageal squamous cell carcinoma).

Adult (16+ years) smoking prevalence between 2003-2007 and second-hand smoking prevalence was collated from Health Survey for England (HSE) datasets (2003-2007) and categorised by sex, 10-year age band and equivalised household income quintiles, accessed through the UK Data Service. A 10-year latency period between smoking exposure and subsequent cancer incidence was used in line with previous methodology.^1,13^ Smoking prevalence for 2004 had to be imputed using a simple linear model based on available years: 2003, 2005, 2006 and 2007.

Relative risk (RR) estimates (see Supplementary Material A) were obtained from meta-analyses through a literature search using previously defined search terms (see Supplementary Material B).^1^ The literature was reviewed between two researchers (NP and KB) to decide on the most appropriate RR estimate to use for each cancer type.

### Population Attributable Fraction formula

To calculate the proportion of cancer cases attributable to smoking by deprivation quintile, the standard population attributable fraction (PAF) formula was used:^13^

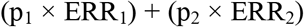

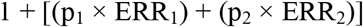

Where p_1_ is the proportion of ‘current cigarette smokers’ in England, p_2_ is the proportion of ‘ex-regular cigarette smokers’, ERR_1_ is the excess relative risk (relative risk – 1) for current smokers and ERR_2_ is the excess relative risk (relative risk – 1) for ex-smokers. Lung cancer had a specific adjustment to the calculation which included an extension of the formula above to account for second-hand smoke exposure prevalence (see Supplementary Material C).

Smoking-attributable cases were calculated for each cancer type and then summed to obtain figures for all smoking-related cancer types combined. Overall PAF estimates used the smoking-attributable cancer cases as the numerator and all cancers combined excluding non-melanoma skin cancer (C00-C97 excl. C44) as the denominator, by sex and deprivation quintile. PAF estimates are presented for all ages combined (0-99+ years) and broken down by two broad age groups (24-64 years and 65+ years). Confidence intervals were not calculated, all comparisons are based on point estimates.

### Observed deprivation gap in cancer incidence and smoking

To further investigate the contribution of smoking to cancer incidence by socio-economic position in England, we grouped smoking-related cancer types together to form a combined age-standardised incidence rate (ASR) by deprivation quintile (2013-2017). We then modelled ASR’s for two hypothetical smoking scenarios based on smoking-related cancer types where: scenario 1) everyone had the same smoking prevalence as the least deprived quintile; scenario 2) nobody smoked. Rates were age-standardised to the 2013 European Standard Population.^14^

To calculate the proportion of avoidable cases under each smoking scenario, we used deprivation-associated cases for deprivation-related cancer types (representing the observed deprivation gap in cancer incidence) as the denominator, and the number of deprivation-associated cases in scenario 1 and scenario 2 as the numerator.

Deprivation-associated cases were calculated using age-specific incidence rates, as has been previously described.^15^ Briefly, ‘expected’ cases were estimated by applying the age-specific incidence rate from the least deprived quintile to each population of the remaining 4 quintiles. The ‘expected’ cases were then subtracted from their corresponding observed cases to produce excess deprivation-associated cases. For the remainder of this article, ‘deprivation-associated cases’ will be used to refer to excess cases due to higher incidence rates in more deprived populations compared to the least deprived.

Confidence intervals were calculated for ASRs, but not for deprivation-associated case estimates. See Supplementary Materials D and E for more detailed information on these calculations.

### Sensitivity analysis

Due to lack of data, the measure of deprivation used for smoking prevalence (equivalised household income) and cancer incidence (income domain of IMD) was not a direct match. To assess the robustness of the main results to differences in deprivation measurement, PAFs were also calculated with smoking prevalence by ‘all domains’ IMD (7 domains: income, employment, health and disability, education, barriers to housing and services, crime and living environment) from HSE datasets (2003-2007), accessed through the UK Data Service.

## Results

### Population Attributable Fraction (PAF) of cancer related to smoking by deprivation quintile

A strong deprivation gradient was observed for the proportion of cancer cases attributable to smoking in England (Table 1). For all ages combined, the smoking PAF was 2.2 times larger in the most deprived quintile compared to the least deprived quintile. The smoking PAF increased from 9.7% in the least deprived quintile to 21.1% in the most deprived quintile.

**Table 1.** Average number and proportion of smoking-attributable cancer cases per year by sex, age and deprivation quintile, England, 2013-2017.

| Deprivation quintile | 25-64 years |  |  | 65+ years |  |  | All ages (0-99+ years) |  |  |
| --- | --- | --- | --- | --- | --- | --- | --- | --- | --- |
|  | Observed cases | PAF* | Smoking attributable cases | Observed cases | PAF* | Smoking attributable cases | Observed cases | PAF* | Smoking attributable cases |
| **Females** |  |  |  |  |  |  |  |  |  |
| **1 (least)** | 11,675 | 4.6% | 542 | 18,654 | 9.7% | 1,817 | 30,626 | 7.7% | 2,359 |
| **2** | 11,919 | 6.3% | 751 | 19,969 | 10.9% | 2,184 | 32,176 | 9.1% | 2,935 |
| **3** | 11,404 | 8.2% | 930 | 18,934 | 13.1% | 2,482 | 30,654 | 11.1% | 3,413 |
| **4** | 11,202 | 10.3% | 1,153 | 17,042 | 16.9% | 2,872 | 28,569 | 14.1% | 4,026 |
| **5 (most)** | 10,990 | 14.3% | 1,566 | 15,009 | 19.9% | 2,991 | 26,398 | 17.3% | 4,558 |
| **Males** |  |  |  |  |  |  |  |  |  |
| **1 (least)** | 8,832 | 9.2% | 810 | 24,049 | 12.6% | 3,031 | 33,203 | 11.6% | 3,841 |
| **2** | 9,067 | 12.1% | 1,099 | 25,223 | 15.1% | 3,812 | 34,616 | 14.2% | 4,911 |
| **3** | 8,954 | 14.9% | 1,332 | 22,743 | 17.5% | 3,979 | 32,011 | 16.6% | 5,311 |
| **4** | 8,945 | 18.1% | 1,619 | 19,768 | 21.4% | 4,227 | 29,059 | 20.1% | 5,846 |
| **5 (most)** | 9,228 | 23.4% | 2,161 | 17,198 | 26.3% | 4,529 | 26,829 | 24.9% | 6,690 |
| **Persons** |  |  |  |  |  |  |  |  |  |
| **1 (least)** | 20,507 | 6.6% | 1,352 | 42,703 | 11.4% | 4,848 | 63,828 | 9.7% | 6,200 |
| **2** | 20,986 | 8.8% | 1,850 | 45,192 | 13.3% | 5,996 | 66,792 | 11.7% | 7,846 |
| **3** | 20,358 | 11.1% | 2,262 | 41,677 | 15.5% | 6,461 | 62,665 | 13.9% | 8,724 |
| **4** | 20,147 | 13.8% | 2,772 | 36,810 | 19.3% | 7,099 | 57,628 | 17.1% | 9,871 |
| **5 (most)** | 20,217 | 18.4% | 3,727 | 32,206 | 23.3% | 7,520 | 53,227 | 21.1% | 11,247 |

\*PAF: Population attributable fraction out of all cancers (excl. non-melanoma skin cancer)

Similar relative increases in PAFs were observed for both sexes, but the PAFs were generally larger for males compared to females. A similar deprivation gradient was found for each broad age group. However, the smoking PAFs were generally smaller in the younger age group compared to the older age group. There was variation in PAFs by cancer type, with both lung cancer and laryngeal cancer having the largest PAFs, as well as strong deprivation gradients (Figures 1a and 1b).

**Figures 1a.**
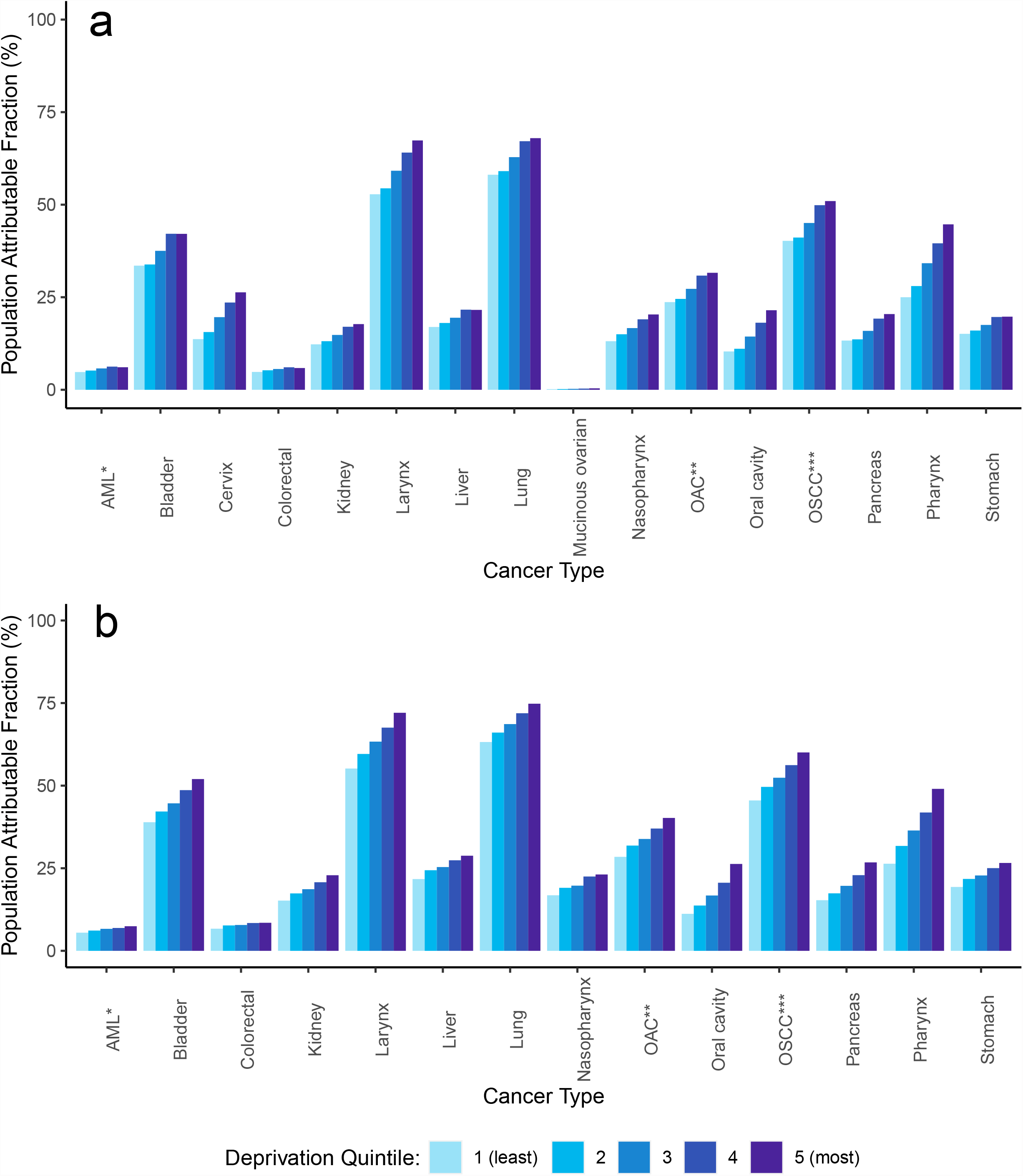
(females) and 1b (males). Population Attributable Fraction (PAF) for smoking, by cancer type and deprivation quintile, England, 2013-2017. *Acute myeloid leukaemia; **Oesophageal adenocarcinoma; ***Oesophageal squamous cell carcinoma

### Cancer incidence by deprivation quintile

Age-standardised incidence rates by deprivation quintile and sex are displayed in Figures 2a and 2b. A clear deprivation gradient is observed for smoking-related cancer types for both sexes, with a 63% and a 60% relative increase in ASR between the least and most deprived quintiles for females and males, respectively.

**Figures 2a.**
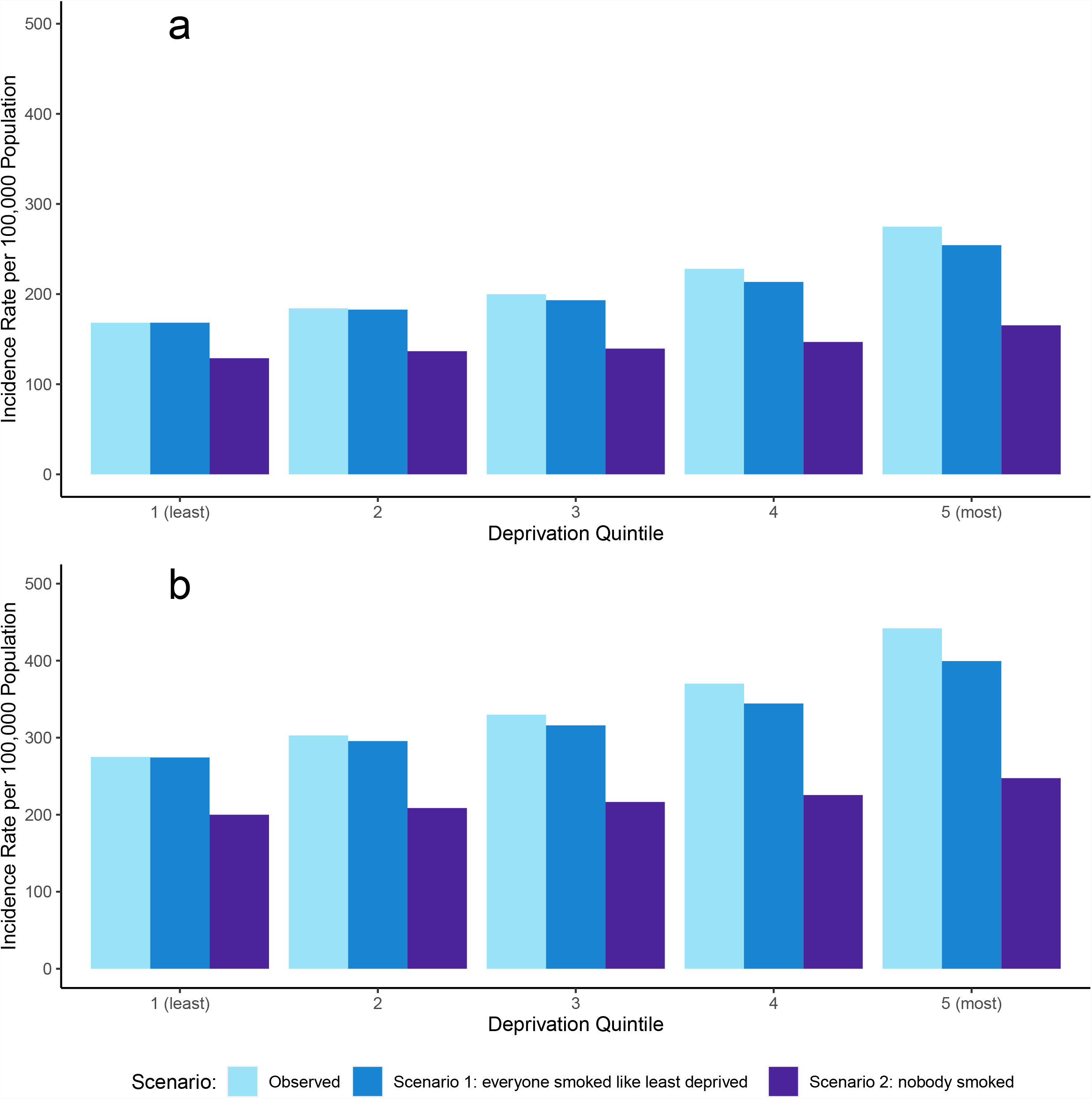
(females) and 2b (males). Combined European Age-Standardised incidence rates (ASR) per 100,000 population for smoking-related cancer types* by deprivation quintile and sex, for observed cancer incidence (the current situation), scenario 1 and scenario 2, England, 2013-2017. *oral cavity, pharynx, nasopharynx, larynx, oesophagus, stomach, colorectal, liver, pancreas, lung, cervix uteri, kidney, bladder, ovarian (mucinous) and leukaemia (acute myeloid)

The deprivation gap for incidence rates between the least and most deprived quintiles is partly reduced in scenario 1 to a 51% and 45% relative increase in ASR between the least and most deprived quintile for females and males, respectively. For scenario 2 where nobody smoked, there is a marked reduction in both the cancer incidence rate and the deprivation gradient, which shows a 28% and 24% relative increase in ASR between the least and most deprived quintile for females and males, respectively.

### Deprivation gap in cancer incidence and smoking

A summary of deprivation-associated cases and the proportion of the observed deprivation gap in cancer incidence that could have been prevented in scenarios 1 and 2 is presented in Table 2. For deprivation-related cancer types, it is estimated that there were an average of 27,156 cases (11,851 in females and 15,305 in males) associated with deprivation every year in England between 2013 and 2017.

**Table 2.** Estimated average number of deprivation-associated cases per year for deprivation-related cancer types* and smoking-related cancer types**, scenario 1 and scenario 2; and the estimated number of deprivation-associated cases and proportion of the observed deprivation gap in cancer incidence that could have been prevented, in England, in 2013-2017.

|  |  | 25-64 years |  |  | 65+ years |  |  | All ages<br>(0-99+ years) |  |  |
| --- | --- | --- | --- | --- | --- | --- | --- | --- | --- | --- |
|  |  | Female | Male | Persons | Female | Male | Persons | Female | Male | Persons |
| **Deprivation-associated cases** | Deprivation - related cancer types | 4,380 | 5,242 | 9,622 | 7,467 | 10,043 | 17,510 | 11,851 | 15,305 | 27,156 |
|  | Smoking-related cancer types | 3,562 | 4,782 | 8,344 | 6,413 | 9,248 | 15,661 | 10,009 | 14,057 | 24,066 |
|  | Scenario 1<sup>1</sup> | 2,605 | 3,481 | 6,086 | 5,403 | 7,012 | 12,415 | 8,043 | 10,519 | 18,562 |
|  | Scenario 2<sup>2</sup> | 1,154 | 1,474 | 2,628 | 2,281 | 2,552 | 4,833 | 3,470 | 4,052 | 7,522 |
| **Preventable deprivation-associated cases (Preventable proportion of the observed deprivation gap in cancer incidence)<sup>a</sup>** | Scenario 1<sup>1</sup> | 957<br>(21.9%) | 1,301<br>(24.8%) | 2,258<br>(23.5%) | 1,010<br>(13.5%) | 2,236<br>(22.3%) | 3,246<br>(18.5%) | 1,966<br>(16.6%) | 3,538<br>(23.1%) | 5,504<br>(20.3%) |
|  | Scenario 2<sup>2</sup> | 2,408<br>(55.0%) | 3,308<br>(63.1%) | 5,716<br>(59.4%) | 4,132<br>(55.3%) | 6,696<br>(66.7%) | 10,828<br>(61.8%) | 6,539<br>(55.2%) | 10,005<br>(65.4%) | 16,544<br>(60.9%) |

<sup>1</sup>Scenario where everyone has the same smoking prevalence as the least deprived quintile; <sup>2</sup>Scenario where nobody smoked
<sup>a</sup>Calculation: $957 = 3562 - 2605$ ; $21.9\% = 957/4380$
\*head and neck (oral cavity, salivary glands, pharynx, nasopharynx, larynx, nasal cavity and middle ear, accessory sinuses), oesophagus, stomach, colorectal, liver, pancreas, lung, cervix uteri, kidney, bladder, small intestine, anal, gallbladder, vulva, vagina, corpus uteri, penis, Hodgkin Lymphoma and cancer of unknown primary
\*\*oral cavity, pharynx, nasopharynx, larynx, oesophagus, stomach, colorectal, liver, pancreas, lung, cervix uteri, kidney, bladder, ovarian (mucinous) and leukaemia (acute myeloid)

If everyone had the same smoking prevalence as the least deprived quintile 20.3% (5,504 cases every year) of deprivation-associated cases could have been prevented. If nobody smoked, 60.9% (16,544 cases every year) of deprivation-associated cases could have been prevented.

### Sensitivity Analysis

The PAFs estimated from smoking prevalence by IMD all domains were similar to the PAFs estimated from smoking prevalence by equivalised household income. For females, the PAFs increased from 7.9% in the least deprived quintile to 18.4% in the most deprived. For males, the PAFs increased from 12.1% in the least deprived to 24.3% in the most deprived (see Supplementary Material F).

## Discussion

### Interpretation of main findings

We observed a strong deprivation gradient for the proportion of cancer cases attributable to smoking in England, which reflects the clear and longstanding socio-economic inequality observed for smoking prevalence in England.^9,16^

Smoking is a key driver of socio-economic inequality in cancer incidence in England. If everyone had the same smoking prevalence as the least deprived quintile 20% (5,504 cases every year) of deprivation-associated cancer cases between 2013 and 2017 could have been prevented. If no one in England had smoked, 61% (16,544 cases every year) of deprivation-associated cases could have been prevented, indicating that smoking explained the majority of the observed deprivation gap in cancer incidence in England between 2013 and 2017.

Though the majority of the observed deprivation gap in cancer incidence can be explained by smoking for both sexes, other risk factors are probably contributing to the remainder of the gap. Obesity (body mass index [BMI] 30+) is positively associated with deprivation for adults in England,^10^ as well as being related to 8 cancer types that are also related to deprivation.^3,8^ Routine and manual workers may have higher risk of exposure to occupational risk factors (e.g. asbestos, silica, aromatic amines) that are related to cancers of the lung, head and neck and bladder.^17,18,19,20^ Prevalence of the human papillomavirus (HPV) infection and helicobacter pylori infection are positively associated with deprivation in the UK, and are linked to numerous cancers that are more common in deprived areas.^8,21,22^

Other research has addressed the hypothetical removal of socio-economic inequality in risk factor exposure on subsequent cancer incidence or mortality, however direct comparisons are precluded by methodological differences (e.g. measure of deprivation, RRs, outcome measures). A French study estimated that 43.4% and 27.5% of deprivation-associated cancer cases for smoking-related cancer types could have been prevented if everyone smoked like the least deprived, in females and males respectively.^11^ In Australia it was estimated that 4% of all cancer cases could have been prevented if smoking, overweight and obesity and physical activity prevalence matched the least deprived across all deprivation quintiles.^12^ Smoking accounted for the vast majority of these deprivation-associated cases. A UK team showed that 30% of lung and laryngeal cancer deaths in men, and 23% of those in women, could be prevented if everyone smoked like those with tertiary education.^23^

### Policy Implications

The UK government’s prevention green paper recently set the aim of England becoming smoke free by 2030, defined as smoking prevalence below 5%.^24^ Successful UK public health initiatives have contributed to overall smoking prevalence declining over time,^25^ but smoking inequalities have widened.^9,16^ The Marmot review of 2010 argued that action is needed across the social gradient ‘with a scale and intensity that is proportionate to the level of disadvantage’.^26^ To incorporate this, action is needed at both a national and local level.

Fiscal measures provide a national cost-effective approach to help target and reduce smoking prevalence, particularly for future generations, whilst also increasing government revenues.^27^ And fiscal measures may also be effective for more deprived smokers where price is more of a potential barrier to consumption.^28,29^ A study modelling the impact of a 10% increase per annum in the price of cigarettes in England and Wales projected a 74% and 86% reduction in the socio-economic gap in lung cancer incidence by 2050, in females and males, respectively.^30^

Local level support can aid smoking cessation for current smokers, particularly for those from the most deprived communities. Smokers from deprived backgrounds are subject to barriers (e.g. lack of social support, high nicotine dependence) that makes it difficult for them to quit.^31,32,33^ Local Stop Smoking Services provide multi-faceted smoking cessation support within communities that can engage with smokers from deprived communities.^33,34^ However, these services are increasingly threatened due to central funding cuts, making it difficult for them provide support locally across the country. Reversing of these cuts would likely help tackle smoking inequalities and prove cost-effective, by reducing smoking-related ill-health that negatively impacts on the National Health Service and productivity.^35,36^

### Strengths and Limitations

We provide a unique quantification of the relationship between socio-economic deprivation, smoking and subsequent cancer incidence in England. Modelling like this may help inform and reinforce policy to prevent smoking-related cancer and improve health more generally in deprived populations. The analysis used high quality cancer incidence and smoking prevalence data, which was averaged over 5 years to reduce the risk of spurious results as a consequence of any year-on-year fluctuation.

This analysis is not without limitations. The same RRs for current and ex-smoking prevalence were applied across all deprivation quintiles. This may reduce the accuracy of the point estimates if the risk associated with those broad definitions varies by deprivation quintile. For example, more deprived smokers may smoke more heavily and start smoking younger.^37,38^ They are also more likely to have multiple cancer risk factors,^39^ including those which combine synergistically with smoking to raise cancer risk, such as alcohol,^40,41^ obesity^42,43^ and occupational exposures.^44,45^ However, the net effect of this is likely to be underestimation of the deprivation gap in smoking PAFs.

These calculations can only be considered estimates because of the PAF methodology used, which is an indirect and relatively simple method that is subject to some uncertainty around point estimates. We used a 10-year latency period, in line with Parkin et al.’s methodology,^13^and this may under-represent the true lag time between smoking exposure and subsequent cancer incidence. A 10-year latency period also assumes people will remain in the same deprivation group from exposure through to recording of cancer incidence. The cancer incidence data uses highly granular area-level rather than individual-level deprivation, meaning these findings may be subject to ecological fallacy.

### Conclusion

Smoking is an important driver of cancer incidence inequalities in England. Efforts to reduce smoking prevalence should focus on minimising smoking inequalities. More research is required to better understand and overcome the complex barriers that smokers from deprived populations face in order to enhance smoking cessation interventions.

## Data Availability

The cancer incidence data analysed during the current study are available from the National Cancer Registration and Analysis Service (part of Public Health England), on request through the Office for Data Release but restrictions apply to the availability of these data, which were used under license for the current study, and so are not publicly available.
The population and smoking prevalence data are publicly available.

https://www.ons.gov.uk/peoplepopulationandcommunity/populationandmigration/populationestimates

https://www.data-archive.ac.uk/

## Additional Information

## Acknowledgements

We acknowledge the work of the England cancer registry, as without their work there would be no incidence data. This work uses data provided by patients and collected by the NHS as part of their care and support. We also acknowledge the work of NHS Digital of the Health Survey for England team at the Health and Social Survey Research group at the Department of Epidemiology at University College London with NatCen Social Research for their smoking prevalence data. Their data provides detailed and reliable breakdowns across a range of different metrics and over many years, that helped enable this research to take place.

## Authors’ contributions

NP carried out the main analysis and wrote the manuscript. KB, IS and JS provided advice and support on the analysis and manuscript. CD provided support by checking methods and data. YK accessed the data and helped prep part of it.

## Ethics approval and consent to participate

Data was de-identified, obtained from either Public Health England via the Office for Data Release or from a publicly available source (UK Data Service). Therefore, no ethical approval was necessary as part of accessing these data.

## Data availability

The cancer incidence data analysed during the current study are available from the National Cancer Registration and Analysis Service (part of Public Health England), on request through the Office for Data Release but restrictions apply to the availability of these data, which were used under license for the current study, and so are not publicly available.

The population and smoking prevalence data are publicly available.

