## Supplementary material for "Socio-economic deprivation and cancer incidence in England: Quantifying the role of smoking"

**Supplementary Material A-F**

Nick W. S. Payne^1*^, Katrina F. Brown^1^, Christine Delon^1^, Yannis Kotrotsios^1^, Isabelle Soerjomataram^2^, Jon Shelton^1^

^1^ Policy, Information and Communication Directorate, Cancer Research UK, 2 Redman Place, London, E20 1JQ

^2^ Section of Cancer Surveillance, International Agency for Research on Cancer, Lyon, France.

Supplementary Material A: ICD-10 codes and relative risks (RRs) for cancer types included in the analysis	2

Supplementary Material B: Search terms for identifying relative risks	4

Supplementary Material C: Population Attributable Fraction formula for lung cancer second-hand smoke exposure	5

Supplementary Material D: Calculation of age-standardised incidence rates (ASR’s) by socio-economic position (deprivation quintile) for scenario 1 and scenario 2 in England	6

Supplementary Material E: Calculation of deprivation-associated cancer cases for deprivation-related cancer types, smoking-related cancer types, scenario 1 and scenario 2 in England	8

Supplementary Material F: Population attributable fractions using all domains IMD for smoking prevalence by sex, age and deprivation quintile	10

References for Supplementary Material A	11

**Supplementary Material A: ICD-10 codes and relative risks (RRs) for cancer types included in the analysis**

| **Cancer type** | **Deprivation-related** | **Smoking-related** | **Excess relative risk** | | | | |
| --- | --- | --- | --- | --- | --- | --- | --- |
|  |  |  | **Current** | | **Former** | | **Source** |
|  |  |  | **Females** | **Males** | **Females** | **Males** |  |
| Oral cavity (C00-C06) | Yes^a^ | Yes | 0.91 | 0.91 | 0.00 | 0.00 | Maasland et al.^[1]^ |
| Salivary Glands (C07-C08) | Yes^a^ | No | - | - | - | - | - |
| Pharynx (C09-C10, C12-C14) | Yes^a^ | Yes | 2.43 | 2.43 | 0.00 | 0.00 | Gandini et al.^[2]^ |
| Nasopharynx (C11) | Yes^a^ | Yes | 0.59 | 0.59 | 0.36 | 0.36 | Long et al.^[3]^ |
| Nasal Cavity and Middle Ear (C30) | Yes^a^ | No | - | - | - | - | - |
| Accessory Sinuses (C31) | Yes^a^ | No | - | - | - | - | - |
| Larynx (C32) | Yes^a^ | Yes | 6.01 | 6.01 | 1.37 | 1.37 | Zuo et al.^[4]^ |
| Oesophageal adenocarcinoma  (C15, ICD-O-3 814-838) | Yes^b^ | Yes | 1.32 | 1.32 | 0.62 | 0.62 | Tramacere et al.^[5]^ |
| Oesophageal squamous cell carcinoma (C15, ICD-O-3 805-808) | Yes^b^ | Yes | 3.21 | 3.21 | 1.18 | 1.18 | Prabhu et al.^[6]^ & Pandeya et al.^[7]^ |
| Stomach (C16) | Yes | Yes | 0.30 | 0.63 | 0.00 | 0.42 | Li et al.^[8]^ |
| Colorectal (C18-C20) | Yes | Yes | 0.11 | 011 | 0.15 | 0.15 | Cheng et al.^[9]^ |
| Liver (C22) | Yes | Yes | 0.66 | 0.66 | 0.51 | 0.51 | Abel-Rahman et al.^[10]^ |
| Pancreas (C25) | Yes | Yes | 0.90 | 0.90 | 0.20 | 0.20 | Lugo et al.^[11]^ |
| Lung (C33-C34) | Yes | Yes | 5.99 | 6.33 | 2.13 | 2.14 | O’Keefe et al.^[12]^ |
| Lung second-hand smoke | - | - | 0.41 | 0.41 | - | - | Jayes et al.^[13]^ |
| Cervix uteri (C53) | Yes | Yes | 1.00 | *-* | 0.00 | *-* | Gandini et al.^2^ |
| Kidney (C64-C66, C68) | Yes | Yes | 0.27 | 0.57 | 0.20 | 0.29 | Liu et al.^[14]^ |
| Bladder (C67) | Yes | Yes | 2.56 | 2.44 | 1.04 | 0.92 | van Osch et al.^[15]^ |
| Small intestine (C17) | Yes | No | *-* | *-* | *-* | *-* | - |
| Anal (C21) | Yes | No | *-* | *-* | *-* | *-* | - |
| Gallbladder (C23) | Yes | No | *-* | *-* | *-* | *-* | - |
| Vulva (C51) | Yes | No | *-* | *-* | *-* | *-* | - |
| Vagina (C52) | Yes | No | *-* | *-* | *-* | *-* | - |
| Uterus (C54-C55) | Yes | No | *-* | *-* | *-* | *-* | - |
| Penis (C60) | Yes | No | *-* | *-* | *-* | *-* | - |
| Cancer of unknown primary  (C77-C80) | Yes | No | *-* | *-* | *-* | *-* | - |
| Hodgkin Lymphoma (C81) | Yes | No | *-* | *-* | *-* | *-* | - |
| Ovary mucinous  (ICD-10 C56-C57.4, ICD-O(3) 8480/3) | No | Yes | 0.44 | *-* | 0.00 | *-* | Santucci et al.^[16]^ |
| Acute myeloid leukaemia  (C920, C924, C925, C926, C928, C930, C940, C942) | No | Yes | 0.52 | 0.52 | 0.45 | 0.45 | Colamesta et al.^[17]^ |

a: Head and neck (C00-C14, C30-C32); b: Oesophagus (C15)

**Supplementary Material B: Search terms for identifying relative risks**

A smoking related search string was combined with smoking-related cancer types using AND. Searches were made in PubMed, and were supplemented using Google Scholar.

| **Risk factor** | **Search string** |
| --- | --- |
| Tobacco | tobacco OR cigarette OR smoking OR environmental tobacco smoke OR secondhand smoke |
| **Cancer Type** |  |
| Kidney cancer | (kidney OR renal OR renal cell) AND (cancer OR carcinoma OR tumour) |
| Bladder cancer | (bladder OR urothelium OR urothelial) AND (cancer OR carcinoma OR tumour) |
| Ovarian cancer | (ovary OR ovarian) AND (cancer OR carcinoma OR tumour) |
| Oral cavity cancer | (oral OR mouth) AND (cancer OR tumour) |
| Nasopharyngeal cancer | (nasopharynx OR nasopharyngeal) AND (cancer OR tumour) |
| Pharyngeal cancer | (oropharynx OR oropharyngeal OR pharynx OR pharyngeal) AND (cancer OR tumour) |
| Oesophageal cancer | (oesophagus OR oesophageal) AND (cancer OR adenocarcinoma OR squamous cell AND (cancer OR tumour) |
| Stomach cancer | (stomach OR gastric OR cardia) AND (cancer OR adenocarcinoma OR tumour) |
| Bowel cancer | (colorectum OR colorectal OR colon OR rectum OR rectal OR bowel) AND (cancer OR tumour) |
| Liver cancer | (liver OR hepatic OR hepatocellular) AND (cancer OR carcinoma OR tumour) |
| Pancreatic cancer | (pancreas OR pancreatic) AND (cancer OR adenocarcinoma OR tumour) |
| Laryngeal cancer | (larynx OR laryngeal) AND (cancer OR tumour) |
| Lung cancer | lung AND (cancer OR adenocarcinoma OR squamous cell carcinoma OR tumour) |
| Leukaemia (acute myeloid) | (leukaemia OR leukemia) AND myeloid |

**Supplementary Material C: Population Attributable Fraction formula for lung cancer second-hand smoke exposure**

(p_1_ × ERR_1_) + (p_2_ × ERR_2_) + (p_3_ × ERR_3_)

----------------------------------------------------------------------

1 + [(p_1_ × ERR_1_) + (p_2_ × ERR_2_) + (p_3_ × ERR_3_)]

Where p_1_ is the proportion of ‘current cigarette smokers’ in England, p_2_ is the proportion of ‘ex-regular cigarette smokers’, p_3_ is the proportion of people exposed to second-hand smoke, ERR_1_ is the excess relative risk (relative risk – 1) for current smokers, ERR_2_ is the excess relative risk (relative risk – 1) for ex-smokers and ERR_3_ is the excess relative risk (relative risk – 1) for second-hand smoke exposure.

**Supplementary Material D: Calculation of age-standardised incidence rates (ASR’s) by socio-economic position (deprivation quintile) for scenario 1 and scenario 2 in England**

Below is a key and corresponding pseudo R code used to calculate ASR’s by deprivation quintile.

**Key:**

CancerCases: number of cancer cases for smoking-related cancer types, scenario 1 and scenario 2 in England (2013-2017). For Scenario 1 and scenario 2, the incidence count was modelled and calculated as follows:

- Scenario 1: a = b-(c-d), whereby:

a: cancer cases for smoking-related cancer types if everyone had the same smoking prevalence as the least deprived

b: observed cancer cases for smoking-related cancer types

c: smoking attributable cancer cases for observed smoking prevalence

d: smoking attributable cancer cases if everyone had the same smoking prevalence as the least deprived

- Scenario 2: x = b – c, whereby:

x: cancer cases for smoking-related cancer types if nobody smoked

b: observed cancer cases for smoking-related cancer types

c: smoking attributable cancer cases for observed smoking prevalence

PopulationCount: population estimates for England (2013-2017).

AgeRange: 5-year age bands for observed smoking related cancer types. 5-year age bands for scenario 1 and scenario 2 between 0-24 years, then 10-year age bands for people aged 25+ because smoking prevalence data is presented in 10-year age bands which dictates the aggregation of the modelled incidence data.

Gender: male or female

IMD_quintile: socio-economic deprivation quintile

PopulationWeighting: 2013 European Standard Population Weight was applied by 5-year age bands for smoking-related cancer types. For scenario 1 and scenario 2, 2013 European Standard Population weight was applied by 5-year age band between 0-24 years, then 10-year age bands for people aged 25+ because smoking prevalence data is presented in 10-year aged bands, which dictates aggregation of the modelled incidence data.

**Code:**

\# Join cancer incidence data with population data and 2013 European Standard Population Weighting for ASR calculation

asrSummary <- inner_join(CancerCases, PopulationCount, by=c("AgeRange", "Gender", "IMD_quintile"))

asrSummary <- inner_join(asrSummary, PopulationWeighting, by="AgeRange")

\# ASR calculation

asrSummary <- asrSummary %>%

mutate(Numerator=ifelse(PopulationCount==0, NA, Cases*100,000 / PopulationCount*PopulationWeighting))

asr_quintile_sex <- asrSummary %>%

group_by(Gender, IMD_quintile) %>%

summarise(ASR = sum(Numerator) / sum(PopulationWeighting)) %>%

ungroup()

**Supplementary Material E: Calculation of deprivation-associated cancer cases for deprivation-related cancer types, smoking-related cancer types, scenario 1 and scenario 2 in England**

Below is a key and corresponding pseudo R code used to calculate the deprivation-associated cases.

**Key:**

CancerCases: number of cancer cases for deprivation- and smoking-related cancer types, scenario 1 and scenario 2 (see appendix 1d for calculation of scenario 1 and scenario 2 cancer cases) in England (2013-2017).

PopulationCount: population estimates for England (2013-2017)

AgeRange: 5-year age bands for observed smoking related cancer types. 5-year age bands for scenario 1 and scenario 2 between 0-24 years, then 10-year age bands for people aged 25+ because smoking prevalence data is presented in 10-year age bands which dictates the aggregation of the modelled incidence data.

Gender: male or female

IMD_quintile: socio-economic deprivation quintile

AgeSpecificRate: age specific incidence rate by 5-year age band for deprivation- and smoking-related cancer types. 5-year age bands for scenario 1 and scenario 2 between 0-24 years, then 10-year age bands for people aged 25+ because smoking prevalence data is presented in 10-year age bands which dictates the aggregation of the modelled incidence data.

DepAssociatedCases: the number of excess cases due to incidence rates being higher in more deprived groups compared to the least deprived in England (2013-2017).

Quintile1_CancerCases: the number of ‘expected’ cancer cases if every deprivation group had the same incidence rate as the least deprived in England (2013-2017).

**Code:**

\# Join data incidence data with population data for calculating rates and deprivation-associated cases

AgeSpecificSummary <- inner_join(CancerCases, PopulationCount, by=c("AgeRange", "Gender", "IMD_quintile"))

\# Age specific rate calculation

AgeSpecificSummary <- AgeSpecificSummary %>%

group_by(Gender, AgeRange, IMD_quintile, PopulationCount) %>%

summarise(AgeSpecificRate = sum(CancerCases) / sum(PopulationCount)) %>%

ungroup()

\# Excess case calculation

DepAssociatedCases <- AgeSpecificSummary %>%

group_by(Gender, AgeRange) %>%

mutate(Quntile1_Rate = AgeSpecificRate[which(IMD_quintile == 1)]) %>%

mutate(Quintile1_CancerCases = Quntile1_Rate * Population) %>%

mutate(ExcessCases = CancerCases – Quintile1_CancerCases %>%

ungroup()

\# Sum excess cases by deprivation quintile

DepAssociatedCases_quintile_sex <- DepAssociatedCases %>%

group_by(Gender, IMD_quintile) %>%

summarise(yearlyExcess = sum(ExcessCases)) %>%

ungroup()

**Supplementary Material F: Population attributable fractions using all domains IMD for smoking prevalence by sex, age and deprivation quintile**

| **Deprivation Quintile** | **25-64 years** | **65+ years** | **All ages (0-99 years)** |
| --- | --- | --- | --- |
| **Females** | | | |
| **1 (least)** | 4.9% | 9.9% | 7.9% |
| **2** | 6.0% | 11.6% | 9.4% |
| **3** | 7.9% | 13.2% | 11.1% |
| **4** | 10.1% | 17.4% | 14.3% |
| **5 (most)** | 14.2% | 22.0% | 18.4% |
| **Males** | | | |
| **1 (least)** | 9.2% | 13.3% | 12.1% |
| **2** | 11.5% | 15.3% | 14.1% |
| **3** | 14.5% | 18.0% | 16.8% |
| **4** | 17.0% | 21.3% | 19.7% |
| **5 (most)** | 23.6% | 25.2% | 24.3% |
